# Handling missing values in the analysis of between-hospital differences in ordinal and dichotomous outcomes: a simulation study

**DOI:** 10.1101/2023.05.30.23290709

**Authors:** Reinier C.A. van Linschoten, Marzyeh Amini, Nikki van Leeuwen, Frank Eijkenaar, Sanne J. den Hartog, Paul Nederkoorn, Jeannette Hofmeijer, Bart J. Emmer, Alida A. Postma, Wim van Zwam, Bob Roozenbeek, Diederik W.J. Dippel, Hester F. Lingsma, MR CLEAN Registry Investigators

**Affiliations:** Department of Public Health, Erasmus MC, Rotterdam, the Netherlands; Department of Gastroenterology & Hepatology, Franciscus Gasthuis & Vlietland, Rotterdam, the Netherlands; Department of Gastroenterology & Hepatology, Erasmus MC, Rotterdam, the Netherlands; Erasmus School of Health Policy & Management, Erasmus University Rotterdam, Rotterdam, the Netherlands ^4^Department of Radiology and Nuclear Medicine, Erasmus MC, Rotterdam, the Netherlands; Department of Neurology, Erasmus MC, Rotterdam, the Netherlands; Department of Neurology, Amsterdam UMC, Amsterdam, the Netherlands; Department of Neurology, Rijnstate Hospital, Arnhem, the Netherlands; Department of Clinical Neurophysiology, University of Twente, Enschede, the Netherlands; Department of Radiology and Nuclear Medicine, Amsterdam UMC, Amsterdam, the Netherlands; Department of Radiology and Nuclear Medicine, MUMC+, Maastricht, the Netherlands; School for Mental Health and Sciences, Maastricht University, Maastricht, the Netherlands

**Author notes:** Corresponding author:* R.C.A. van Linschoten, Franciscus Gasthuis & Vlietland, P.O Box 10900, 3004BA, Rotterdam, Netherlands.

## Abstract

Missing data are frequently encountered in registries that are used to compare performance across hospitals. The most appropriate method for handling missing data when analysing differences in outcomes between hospitals is unclear. We aimed to compare methods for handling missing data when comparing hospitals on ordinal and dichotomous outcomes. We performed a simulation study using data came from the MR CLEAN registry, a prospective cohort study in 17 hospitals performing endovascular therapy for ischemic stroke in the Netherlands. The investigated methods for handling missing data, both case-mix adjustment variables and outcomes, were complete case analysis (CCA), single imputation, multiple imputation, single imputation with deletion of imputed outcomes and multiple imputation with deletion of imputed outcomes. Data were generated as missing completely at random (MCAR), missing at random (MAR), and missing not at random (MNAR) in three scenarios: (1) 10% missing data in case-mix and outcome; (2) 40% missing data in case-mix and outcome; and (3) 40% missing data in case-mix and outcome with varying degree of missing data among hospitals. Validity and reliability of the methods were compared on the mean squared error (MSE, a summary measure combining bias and precision) relative to the centre effect estimates from the complete reference dataset. For both the ordinal outcome (i.e. the modified Rankin scale) and a common dichotomized version thereof, the MSE of all methods was on average lowest under MCAR and with fewer missing data, and highest with more missing data and under MNAR. The ‘multiple imputation, then deletion’ method had the lowest MSE for both outcomes under all simulated patterns of missing data. Thus, when estimating centre effects on ordinal and dichotomous outcomes in the presence of missing data, the least biased and most precise method to handle these missing data is ‘multiple imputation, then deletion’.

## BACKGROUND

Benchmarking of hospital performance is a promising tool to improve healthcare, but a first requirement is comprehensive, accurate, and complete data.[1, 2] Although a lot of effort is being put into collecting complete data, even well-designed registries suffer from missing data.[3, 4] Missing data can occur in the outcomes of interest, but also in patient characteristics that are used for adjustment, so called case-mix variables. When measuring hospital performance by estimating hospital effects on outcome in registry data, missing data can introduce bias and reduce precision in several ways.[5–7] First, hospital effects that are estimated from smaller samples will have larger variance. Consequently, hospitals with a large proportion of missing data are less likely to be identified as significantly better or worse than the average. Second, patients with complete information tend to be systematically different from patients with missing data,[8] which can reduce validity of the hospital effect estimates. Third, the reason why data are missing may be related to specific characteristics of the hospitals being compared, for example with respect to data collection arrangements.[9] Consequently, missing data might confound between-hospitals comparisons on outcome.

Generally speaking, three missing data mechanisms can be distinguished: missing completely at random (MCAR), missing at random (MAR), and missing not at random (MNAR). Under MCAR, values are missing in a random subset of patients. When missing data are related to other observed patient data, missing data are MAR. For example, data would be MAR when women are more likely to have missing data than men. When missing data are related to unobserved data such as the value of the missing data itself, data are MNAR.[10, 11] An example of data being MNAR would be patients in poorer health not responding to a quality of life questionnaire.

Several methods for handling missing data have been described.[12–15] Complete case analysis (CCA), single imputation, and multiple imputation are techniques that are frequently used. These methods are computationally straightforward, versatile, relatively easy to apply, and available in standard statistical software programs.[12] CCA is a deletion-based method, in which observations with any missing value are excluded from the analysis. With single imputation, a regression model with the variable of interest (i.e., the one with missing value(s)) as dependent variable and patient, treatment, and hospital characteristics as independent variables is fitted. Missing values are then imputed using the value as predicted by the model. With single imputation the uncertainty around the predictions generated by the regression is ignored in the further analysis; the imputed data are analysed as they were observed. Multiple imputation is similar to single imputation in that a regression model is used to model missing data. This method differs from single imputation in that it generates more than one dataset with slightly different, yet plausible, values for the missing observations. These datasets can separately be analysed and the results pooled. In this way multiple imputation does account for uncertainty around the predictions generated by the regression model for missing values.[15] An extension of single and multiple imputation is a method called ‘imputation, then deletion’. With this method, data are first imputed by single or multiple imputation. Then, the imputed outcomes are deleted from the imputed datasets and patients who initially had missing outcome data are not included in the analysis. Imputed values in the case-mix variables are kept. This can improve efficiency compared to multiple imputation, and is more robust to misspecification of the imputation regression model.[16]

Despite missing data often being a problem in between-hospital comparisons, no recommendations exist on which of the aforementioned methods to handle missing data to use when estimating hospital effects. In this simulation study, we aimed to assess the validity and reliability of these methods in estimating between-hospital differences in outcome.

## METHODS

### Study population

Simulations were based on the observational data from the MR CLEAN Registry, of which the methodology has been published.[17] In brief, this large nationwide stroke registry is a prospective observational cohort study in all seventeen hospitals that perform endovascular therapy (EVT) for acute ischemic stroke in the Netherlands. For the registry extensive clinical and neuro-imaging data are collected. We aimed to compare hospital effects on outcome based on different methods of handling missing data with those from a complete reference dataset.

Therefore, we selected patients from the registry with complete data for the variables in the model described below.

### Variables

Our simulation involved the analysis of an ordinal and dichotomous outcome. The ordinal outcome variable was the modified Rankin Scale (mRS).[18] The mRS is a commonly used measure of patients’ functional outcome after ischemic stroke and ranges from 0 (no symptoms) to 6 (death). The dichotomous outcome variable was good functional outcome, defined as an mRS of 0 to 2.[19] The mRS was assessed at 90 days after EVT (± 14 days). For case-mix adjustment, we used the following baseline variables: age, sex, and the National Institute of Health Stroke Scale (NIHSS). The NIHSS measures stroke-related neurologic deficits and ranges from 0 (no stroke symptoms) to 42 (severe stroke). To improve model stability, age and NIHSS were standardised. We also used two process measures as auxiliary variables in the imputation procedure. Auxiliary variables are variables that contain information about the incomplete variables and can improve imputation.[16, 20] These variables were time from onset to groin puncture and the use of general anaesthesia during the EVT procedure as these are likely related to the outcome.[21]

### Statistical analysis

The analysis consisted of three steps (Figure 1). In the first step, missing data were simulated assuming three different mechanisms of missing data: MCAR, MAR and MNAR. Multivariate missing data were simulated using the mice package in R.[22] Details of the procedure are described in the Supplementary Methods. In addition, we simulated three different scenarios for the degree of missing data. In scenario 1, 10% of data on both case-mix (age, sex, and baseline NIHSS) and outcome (ordinal and dichotomous mRS) was made missing in all 17 hospitals. In scenario 2, we assumed 40% of missing data in the outcome and in one important case-mix variable (NIHSS) at baseline, equally distributed over all hospitals. Scenario 3 was the same as scenario 2, but with the probability of data being missing varying between hospitals. The relatively high proportion of 40% missing data per variable in scenario 2 and 3 was based on findings from Dutch and American quality registries for acute stroke care.[3, 4]

**Figure 1.**
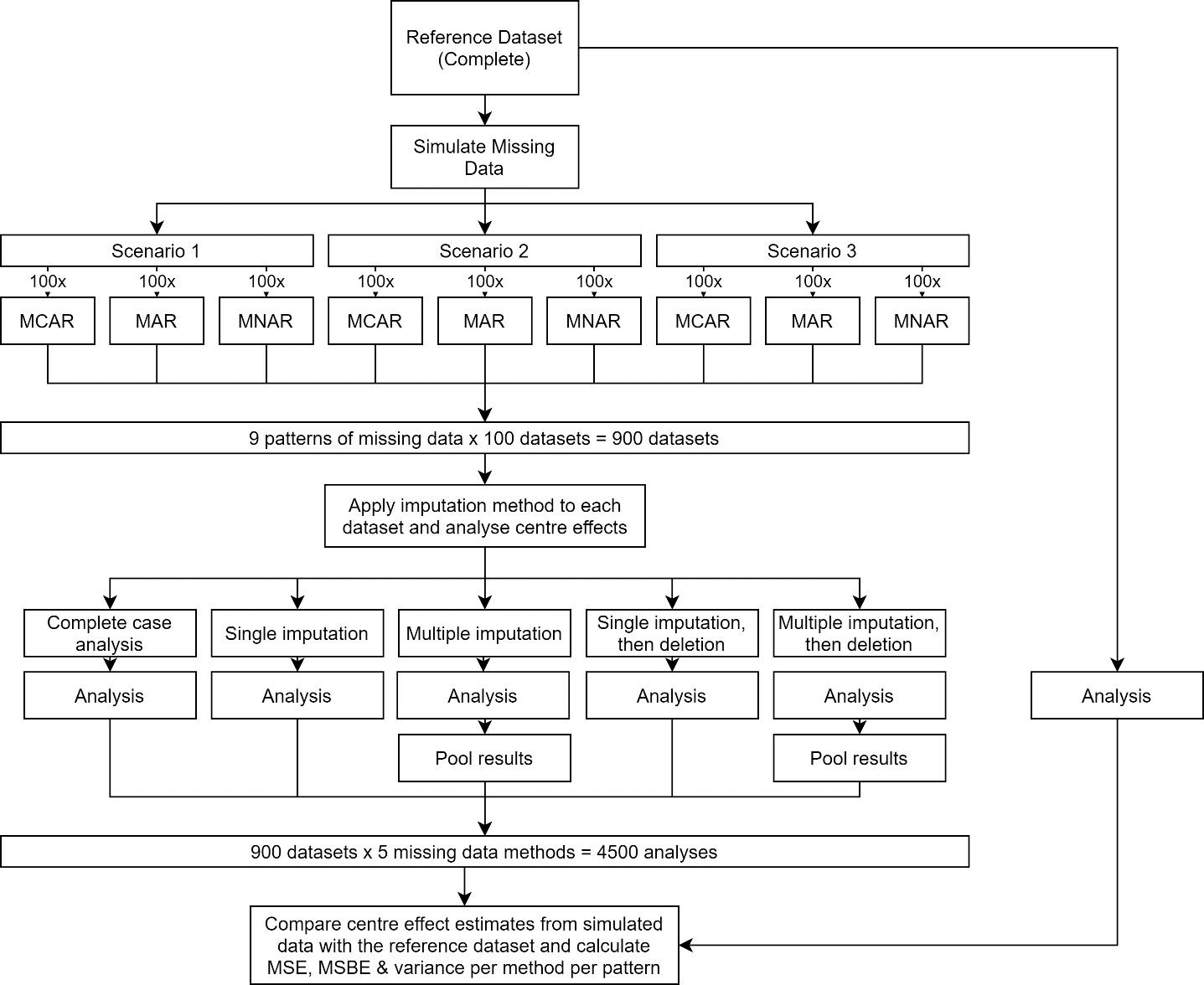
Flowchart of the study methods MCAR: Missing completely at random MAR: Missing at random MNAR: Missing not at random MSE: Mean squared error MSBE: Mean squared bias error Scenario 1: 10% of data missing in both case-mix (age, sex, and baseline NIHSS) and outcome (mRS) in all 17 hospitals. Scenario 2: 40% of data missing in the outcome and baseline NIHSS equally distributed over all hospitals. Scenario 3: 40% of data missing in the outcome and baseline NIHSS with the probability of missing data varying between hospitals.

Within each scenario, we simulated cases with different combinations of missing data (i.e., a patient could have missing data in the outcome, the NIHSS or in both for scenario 2). Applying these combinations means that the proportion of patients with missing data was higher than the proportion of missing data per variable. For example, in scenario 2 (with 40% of missing data per variable), 20% of patients had missing data in only the outcome, 20% in only the NIHSS, and 20% in both the NIHSS and outcome. The percentage of patients with missing data was 20% in scenario 1 and 60% in both scenario 2 and 3.

Combining the three mechanisms and three scenarios led to nine patterns of missing data. To account for random variation, each combination was simulated 100 times.

In the second step, we applied five methods to handle missing data. These were complete case analysis (CCA), single imputation (SI), multiple imputation (MI), ‘single imputation, then deletion’ (SID) and ‘multiple imputation, then deletion’ (MID). Imputation was performed with the mice package in R.[22] A binomial generalised linear mixed model was used to impute the dichotomous outcome, while predictive mean matching based on a linear mixed model was used to impute the ordinal outcome. For further details on the imputation procedure, see the Supplementary Methods.

In the final step, generalised linear mixed models were used to estimate between-hospital differences in outcome for each imputation method (Figure 1). A model with fixed effects for age, sex and baseline NIHSS, a random effect for hospital, and the mRS as the dependent variable was fitted, separately for the ordinal and dichotomous version. For the ordinal outcome, we used proportional odds regression modelling with the mRS reversed, so that a positive fixed or random effect indicates higher odds of a better outcome. For the dichotomous outcome, a binomial generalised linear mixed model was used. Estimates from the multiple imputed datasets were pooled using Rubin’s rules.[15]

The hospital effect estimates from these models were used to assess the validity and reliability of the five methods for handling missing data across the nine patterns of missing data (three scenarios times three mechanisms of missing data). Validity was measured by the mean squared bias error (MSBE), which is the squared mean difference between the estimates of hospital effects for each simulated dataset and the hospital effect estimates from the reference dataset[23]. The MSBE can be interpreted as the average squared distance between the estimated random effect and the random effect from the reference dataset. A higher MSBE implies lower validity. Reliability was defined as the variance of hospital effect estimates from the simulated data around the mean random effect from the simulated data. This can be interpreted as the spread of random effects, with a higher variance indicating lower reliability.

Together, MSBE and variance sum to mean squared error (MSE), defined as the mean of the squared differences between hospital effect estimates from the simulated dataset and the hospital effect estimates from the reference dataset.[23] MSBE, variance and MSE were all calculated on the linear predictor scale. When an estimator is unbiased, the MSBE is zero and the MSE equals the variance of the estimator.

## RESULTS

A total of 2817 patients were included in the reference dataset (Supplementary Table 1). Mean patient age ranged from 66 to 75 years and differed among hospitals (p=0.0018). The percentage of female patients did not vary significantly across hospitals (range 44%-67%, p=0.82). Hospitals also differed in mean baseline NIHSS score (13-17, p<0.0001), mean time to groin (183-253, p<0.0001) and use of general anaesthesia (0%-99%, p=0.0005). Outcome differed between hospitals; the percentage of patients with a good functional outcome varied from 35%-51% (p<0.0015). The odds ratios for the reference centre effect estimates ranged from 0.71 to 1.65 for the ordinal outcome and from 0.70 to 1.79 for the dichotomous outcome.

The MSE ranged between 0.0006 and 0.16 for the ordinal outcome and between 0.0007 and 0.20 for the dichotomous outcome (Figure 2 and 3 and Supplementary Tables 2 and 3 for details). The MSE was on average lowest in scenario 1 and under MCAR, and highest in scenario 3 and under MNAR. Overall, the ‘imputation, then deletion’ methods performed best for both outcomes, in all scenarios, and under all mechanisms of missing data. MID slightly outperformed SID, while the standard imputation methods were almost always outperformed by CCA. Only in scenario 3 under MNAR did the standard imputation methods outperform CCA when analysing an ordinal outcome.

**Figure 2.**
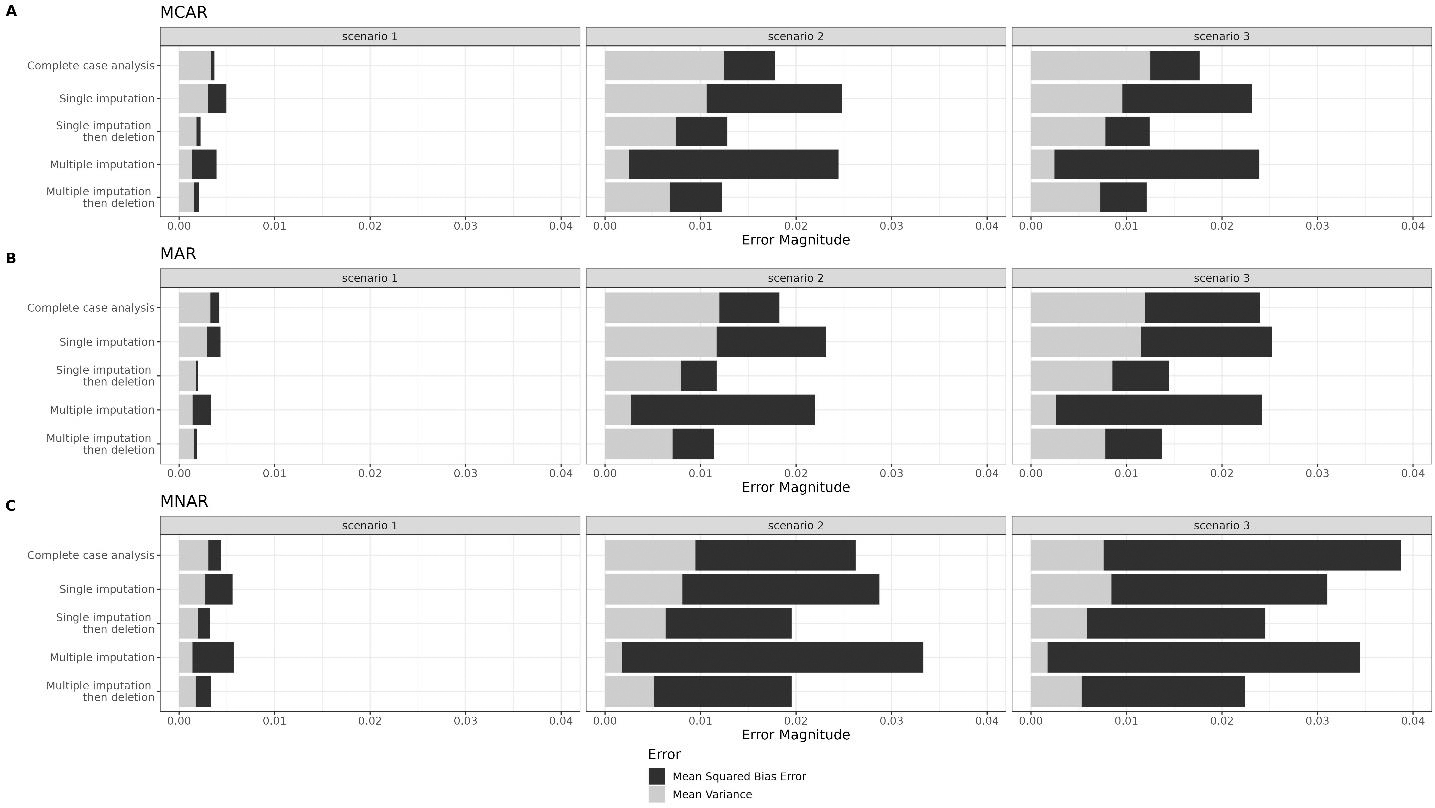
Mean squared error, mean squared bias error and mean variance per method for each scenario under (A) MCAR, (B) MAR, and (C) MNAR for an ordinal outcome MCAR: Missing completely at random MAR: Missing at random MNAR: Missing not at random Scenario 1: 10% of data missing in both case-mix (age, sex, and baseline NIHSS) and outcome (mRS) in all 17 hospitals. Scenario 2: 40% of data missing in the outcome and baseline NIHSS equally distributed over all hospitals. Scenario 3: 40% of data missing in the outcome and baseline NIHSS with the probability of missing data varying between hospitals.

**Figure 3.**
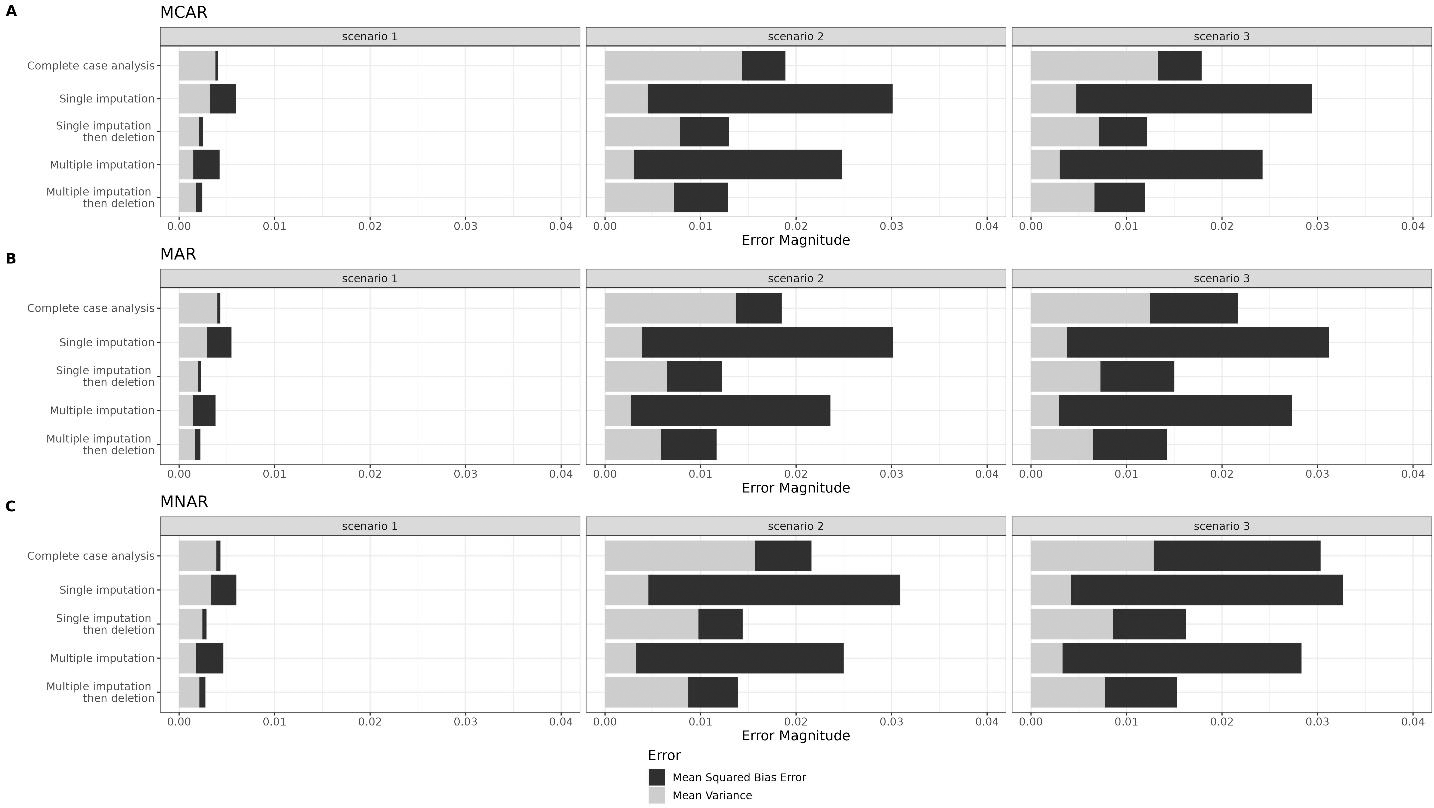
Mean squared error, mean squared bias error and mean variance per method for each scenario under (A) MCAR, (B) MAR, and (C) MNAR for a dichotomous outcome MCAR: Missing completely at random MAR: Missing at random MNAR: Missing not at random Scenario 1: 10% of data missing in both case-mix (age, sex, and baseline NIHSS) and outcome (mRS) in all 17 hospitals. Scenario 2: 40% of data missing in the outcome and baseline NIHSS equally distributed over all hospitals. Scenario 3: 40% of data missing in the outcome and baseline NIHSS with the probability of missing data varying between hospitals.

For CCA, MID and SID, the MSE was mostly driven by variance under MCAR and MAR. By contrast, for SI and MI under MCAR and MAR, bias (i.e., MSBE) was the main driver of the MSE, more so for MI than for SI. Under MNAR, the MSE of all methods was mostly driven by bias.

Centres with larger absolute reference effects had larger MSE for both outcomes, under all methods, mechanisms and scenarios (Supplementary Figures 1 and 2). These results apply to both the ordinal and dichotomous outcome.

## DISCUSSION

In this study, we used registry data to compare the validity and reliability of different methods of handling missing data when estimating centre effects on outcome, assuming nine different patterns of missing data. We found that across these scenario’s, the ‘imputation, then deletion’ methods results in the least biased estimates of centre effects, for both for ordinal and dichotomous outcomes.

Theoretically, under certain assumptions, both the CCA and imputation methods can result in valid estimates for coefficients in regression models. CCA is valid if maximum likelihood approaches are used (as in generalised linear mixed models), data are MCAR or MAR, and we condition our analysis on variables that govern missingness.[24, 25] Imputation methods are valid if missing data are MCAR or MAR and if the imputation model is properly specified. This means that the imputation model should contain the same variables, interactions and nonlinearities as the regression model in the main analysis (in our case the hospital effect estimation), as well as variables that govern missingness.[26] However, after SI, standard errors (and consequently confidence intervals and statistical inference) are invalid, as this method does not properly take into account uncertainty around the missing values. As a result, the standard errors are underestimated. By contrast, MI does properly account for the uncertainty and leads to valid standard errors.[25] In our simulation study we found bias for each method to handle missing values, under every scenario and mechanism of missingness. These results can likely be explained by three reasons. First, while CCA is valid under MAR and MCAR and we properly specified the analysis model (the analysis was conditioned on the variables that governed missingness), mixed models apply shrinkage to the estimated random effects. Shrinkage improves estimation of the random effects by pulling them towards the mean, with the degree of shrinkage being influenced by the ratio of centre-level variability versus residual variability, sample size per centre and effect size.[27, 28] Specifically, shrinkage is lower with higher variability on the centre level, and centres with larger effects sizes and smaller sample sizes are shrunk more. As missing data reduces the sample size that can be used in the analysis, shrinkage will be larger and random effects biased. The effect of reduced sample size can also be seen with the increase in error of CCA from scenario 1 (20% of patients with missing data) to scenario 2 and 3 (60% of patients with missing data).

Second, comparing the results from CCA with the standard imputation methods SI and MI, we would assume centre effect estimates from imputation to be unbiased, as sample sizes are equal to the reference dataset. However, error from the imputation methods is almost always larger than with CCA, and consists mostly of bias. This result could be explained by problematic imputations of the outcome.[16] In our imputation model, we used predictive mean matching to impute the ordinal outcome, because to our knowledge there is no imputation model that can handle clustered ordinal data. Predictive mean matching calculates a predicted value for the outcome from a linear mixed model, and then samples the outcome from a selected set of donors (patients without missing outcomes) with comparable predicted values. The imputation method used in this paper does not allow for drawing donors from only the same centre. As the outcome can therefore be sampled from a different centre, outcomes from centres will resemble each other more, leading to less centre-level variability, increased shrinkage and thus more bias. However, the same results were found for the dichotomous outcome, for which the imputation model was correctly specified: a binomial generalised linear mixed model was used and all variables that govern missingness were included. A possible explanation is that shrinkage is also applied in the generalised linear mixed models used in the imputation procedure. This means that the imputed outcomes are shrunk towards the mean for each centre, again leading to centre effects that resemble each other more with less centre-level variability and thus more bias as a result.

Third, the bias under MNAR could be expected, as none of the methods for handling missing data is theoretically valid under MNAR. The possibility of data being MNAR should always be considered and, if suspected, sensitivity analyses should be performed.[29]

In this study, the least biased method for the estimated centre effects on outcome is ‘imputation, then deletion’. This method increase sample size by including patients with missing case-mix variables, while preventing the problematic imputations of the outcome. Although we did not test the precision of the estimates (i.e. the variance around the point estimates of centre effects), from theory we know that the single imputation methods are invalid and the ‘multiple imputation, then deletion’ approach would be preferred.[25]

Previous research has primarily focused on valid estimation of fixed effects in situations with missing data.[16, 25] One study has looked at the bias of different multiple imputation methods when analysing incomplete data with a linear mixed effects model, and found that these validly estimated the variance of the random intercept distribution.[30] However, this study did not assess the validity and reliability of the random effects themselves, and only considered linear mixed models. We add to this literature by showing that random effects do not behave the same as fixed effects due to shrinkage in the estimation procedure for both ordinal and dichotomous outcomes.

However, some limitations of our research should be noted. First, this is a simulation study based on one dataset. As such, results may not be directly generalizable to other settings, such as when outcomes or random effects have other distributions than those we have studied.

While we aimed to provide a theoretical underpinning of our results, future studies should further investigate random effect estimation when data are missing. Second, we have only assessed the validity and reliability of the point estimates of the centre effects. Precision of the centre effects, as measured with standard errors and coverage of confidence intervals, was not evaluated, but is clearly also important when benchmarking hospitals.[31] Multiple imputation is likely less efficient than maximum likelihood methods, which is due to the fact that the imputed outcomes vary between imputed datasets, which reduces precision.[15] Therefore, future studies should also evaluate precision.

In conclusion, in case of missing data, estimates of centre effects on ordinal and dichotomous outcomes are nearly always biased. The size of the bias is influenced by the proportion and mechanism of missing data. Reliability of the centre effect estimates depend considerably on the method of imputation. The ‘multiple imputation, then deletion’ method seems the most promising method for handling missing data in terms of both validity and reliability.

## FUNDING

The MR CLEAN Registry was partly funded by TWIN Foundation, Erasmus MC University Medical Centre, Maastricht University Medical Centre, and Amsterdam UMC.

## DISCLOSURES

WvZ has received consulting fees from Codman and Stryker DD has received research grants from Stryker and Bracco Imaging The other authors have nothing to disclose.

## DATA SHARING

Data may be obtained from a third party and are not publicly available. The data of the study cannot be made available to other researchers, as Dutch law prohibits data sharing when no patient approval was obtained for sharing coded data. However, syntax or output files of the statistical analyses may be made available for academic purposes upon reasonable request.

## AUTHOR CONTRIBUTION

RCAvL, MA, NvL and FE designed the study. RCAvl and MA undertook analyses and interpretation of study findings and wrote the first draft of the manuscript. NvL, FE and HL contributed to the interpretation of study findings, reviews, and revision of the manuscript. All the authors critically reviewed the various versions of the full paper and approved the final manuscript for submission.

## ETHICAL APPROVAL

The MR CLEAN Registry was approved by the ethics committee of the Erasmus University MC, Rotterdam, The Netherlands (MEC-2014-235). With this approval it was approved by the research board of each participating center. At UMC Utrecht, approval to participate in the study has been obtained from their own research board and ethics committee.

## Notes

### Author Declarations

The ethics committee of the Erasmus University MC, Rotterdam, The Netherlands (MEC-2014-235) gave ethical approval for the MR CLEAN registry.

