## Supplementary File for "Handling missing values in the analysis of between-hospital differences in ordinal and dichotomous outcomes: a simulation study"

### SUPPLEMENTARY INFORMATION

#### Supplementary Methods

*Multivariate amputation procedure*

Datasets with missing data were simulated with the ampute function from the mice package in R.[1] The necessary input of this function is the data, the desired proportion of missing data, the mechanism of missing data and the variables that influence whether data is missing. For scenario 1 we specified that each case-mix variable should have 10% missing data, with patterns where patients could have missing data in one, two or all three of the variables. Under MCAR, data was missing made at random. Under MAR, data was made missing dependent on the case-mix variables that were not missing in the pattern, as well as the outcome. Under MNAR, data was made missing dependent on its own value, as well as the value of the other variables that were made missing in the pattern.

For scenario 2, we specified that the NIHSS and the MRS should have 40% missing data, with patterns where patients could have missing data in either or in both variables. Under MCAR, data was missing made at random. Under MAR, data was made missing dependent on the variables that were not missing in the pattern. Under MNAR, data was made missing dependent on its own value, as well as the value of the other variables that were made missing in the pattern. For scenario 3, the patterns and dependencies were kept equal except that under MAR and MNAR, probability of data being missing was also depended on the hospital.

*Imputation procedure*

Missing data were imputed using the mice package in R.[1] When imputing data, regression models are made for each variable with missing data. Predictors in these models should be variables that are related to the likelihood that data is missing or to the value of missing data.[2] There also needs to be congeniality between the imputation and analysis models, which means that the imputation model contains at least the same variables and interactions as the analysis model[3]. In single imputation, the predicted value from the regression model is used to fill in the missing data point. In multiple imputation the uncertainty around the missing data point is taken into account. This is done by not using the coefficients from the regression model, but by sampling these coefficients from a normal distribution based on the coefficients and their standard errors from the model. These are then used to predict the missing value, but instead of using the predicted value, we sample a value from the prediction interval. These two steps properly account for the uncertainty around the missing data.[2]

In our case, we used age, sex, baseline NIHSS, mRS, as fixed effects and hospital as random effect to impute missing values to ensure congeniality and because these variables were related to probability of data being missing. We added time from stroke onset to groin puncture and use of general anaesthesia as auxiliary variables, as these are related to the outcome and could improve imputation.[4] A binomial generalised linear mixed model was used to impute the dichotomous outcome, while predictive mean matching based on a linear mixed model was used to impute missing data for all variables where single imputation used the predicted values (type 0) and multiple imputation used random draws from the distribution of the coefficients (type 1). For single imputation one dataset was imputed. For multiple imputation approach, we imputed 10 datasets to take into account uncertainty around the missing data.[2]

#### Supplementary Tables

**Supplementary Table 1.** Baseline characteristics of the complete dataset.

| Centre | Female[1] | Age[2] | NIHSS[2] | Time to groin (minutes) [2] | General anaesthesia[3] | mRS 0-2[1] |
|---|---|---|---|---|---|---|
| Centre 01, N = 359 | 172 (48%) | 68 (14) | 16 (6) | 216 (84) | 78 (22%) | 141 (39%) |
| Centre 02, N = 305 | 141 (46%) | 71 (14) | 16 (6) | 213 (83) | 11 (3.6%) | 114 (37%) |
| Centre 03, N = 229 | 119 (52%) | 72 (12) | 14 (6) | 224 (99) | 44 (19%) | 87 (38%) |
| Centre 04, N = 277 | 129 (47%) | 70 (14) | 15 (7) | 224 (89) | 20 (7.2%) | 97 (35%) |
| Centre 05, N = 59 | 30 (51%) | 66 (16) | 16 (6) | 201 (96) | 14 (24%) | 22 (37%) |
| Centre 06, N = 91 | 47 (52%) | 73 (15) | 16 (7) | 191 (85) | 1 (1.1%) | 36 (40%) |
| Centre 07, N = 72 | 34 (47%) | 70 (12) | 14 (6) | 199 (80) | 7 (9.7%) | 36 (50%) |
| Centre 08, N = 108 | 47 (44%) | 71 (13) | 13 (6) | 183 (103) | 35 (32%) | 48 (44%) |
| Centre 09, N = 215 | 108 (50%) | 68 (15) | 15 (6) | 253 (190) | 4 (1.9%) | 75 (35%) |
| Centre 10, N = 94 | 52 (55%) | 69 (15) | 17 (7) | 213 (111) | 11 (12%) | 45 (48%) |
| Centre 11, N = 18 | 12 (67%) | 75 (13) | 14 (4) | 202 (91) | 0 (0%) | 8 (44%) |
| Centre 12, N = 197 | 96 (49%) | 72 (13) | 17 (6) | 195 (81) | 1 (0.5%) | 100 (51%) |
| Centre 13, N = 267 | 121 (45%) | 70 (14) | 15 (5) | 241 (90) | 258 (97%) | 136 (51%) |
| Centre 14, N = 162 | 74 (46%) | 69 (15) | 15 (6) | 205 (79) | 160 (99%) | 65 (40%) |
| Centre 15, N = 57 | 31 (54%) | 72 (13) | 16 (7) | 207 (93) | 8 (14%) | 18 (32%) |
| Centre 16, N = 136 | 63 (46%) | 71 (14) | 15 (6) | 185 (84) | 10 (7.4%) | 51 (38%) |
| Centre 17, N = 181 | 87 (48%) | 71 (14) | 16 (6) | 201 (104) | 4 (2.2%) | 68 (38%) |
| p-value for between-centre differences | 0.82 | 0.0018 | <0.0001 | <0.0001 | 0.0005 | 0.0015 |

NIHSS: National Institute of Health Stroke Scale
mRS: modified Rankin Scale
[1]n (%), Pearson's Chi-squared test
[2]Mean (SD), One-way ANOVA
[3]n (%), Fisher's Exact Test for Count Data with simulated p-value (based on 2000 replicates)

**Supplementary Table 2.** Results from the methods for handling missing data in an ordinal outcome

MCAR: Missing completely at random

MAR: Missing at random

MNAR: Missing not at random

Scenario 1: 10% of data missing in both case-mix (age, sex, and baseline NIHSS) and outcome (mRS) in all 17 hospitals.

Scenario 2: 40% of data missing in the outcome and baseline NIHSS equally distributed over all hospitals.

Scenario 3: 40% of data missing in the outcome and baseline NIHSS with the probability of missing data varying between hospitals.

| Method | Mean Variance | Mean Squared Bias Error | Mean Squared Error |
|---|---|---|---|
| **MCAR - scenario 1** | | | |
| Complete case analysis | 0.0033 | 0.00035 | 0.0037 |
| Single imputation | 0.0030 | 0.0019 | 0.0050 |
| Single imputation then deletion | 0.0018 | 0.00043 | 0.0022 |
| Multiple imputation | 0.0014 | 0.0026 | 0.0039 |
| Multiple imputation then deletion | 0.0016 | 0.00054 | 0.0021 |
| **MAR - scenario 1** | | | |
| Complete case analysis | 0.0033 | 0.00093 | 0.0042 |
| Single imputation | 0.0029 | 0.0014 | 0.0043 |
| Single imputation then deletion | 0.0018 | 0.00024 | 0.0020 |
| Multiple imputation | 0.0014 | 0.0019 | 0.0034 |
| Multiple imputation then deletion | 0.0016 | 0.00032 | 0.0019 |
| **MNAR - scenario 1** | | | |
| Complete case analysis | 0.0031 | 0.0013 | 0.0044 |
| Single imputation | 0.0027 | 0.0029 | 0.0056 |
| Single imputation then deletion | 0.0020 | 0.0013 | 0.0032 |
| Multiple imputation | 0.0014 | 0.0043 | 0.0057 |
| Multiple imputation then deletion | 0.0017 | 0.0016 | 0.0033 |
| **MCAR - scenario 2** | | | |
| Complete case analysis | 0.012 | 0.0053 | 0.018 |
| Single imputation | 0.011 | 0.014 | 0.025 |
| Single imputation then deletion | 0.0074 | 0.0053 | 0.013 |
| Multiple imputation | 0.0025 | 0.022 | 0.024 |
| Multiple imputation then deletion | 0.0068 | 0.0054 | 0.012 |

| | | | |
|---|---|---|---|
| **MAR - scenario 2** | | | |
| Complete case analysis | 0.012 | 0.0063 | 0.018 |
| Single imputation | 0.012 | 0.011 | 0.023 |
| Single imputation then deletion | 0.0079 | 0.0037 | 0.012 |
| Multiple imputation | 0.0028 | 0.019 | 0.022 |
| Multiple imputation then deletion | 0.0071 | 0.0044 | 0.011 |
| **MNAR - scenario 2** | | | |
| Complete case analysis | 0.0095 | 0.017 | 0.026 |
| Single imputation | 0.0081 | 0.021 | 0.029 |
| Single imputation then deletion | 0.0063 | 0.013 | 0.020 |
| Multiple imputation | 0.0018 | 0.032 | 0.033 |
| Multiple imputation then deletion | 0.0051 | 0.014 | 0.020 |
| **MCAR - scenario 3** | | | |
| Complete case analysis | 0.012 | 0.0052 | 0.018 |
| Single imputation | 0.0096 | 0.014 | 0.023 |
| Single imputation then deletion | 0.0078 | 0.0046 | 0.012 |
| Multiple imputation | 0.0025 | 0.021 | 0.024 |
| Multiple imputation then deletion | 0.0073 | 0.0049 | 0.012 |
| **MAR - scenario 3** | | | |
| Complete case analysis | 0.012 | 0.012 | 0.024 |
| Single imputation | 0.012 | 0.014 | 0.025 |
| Single imputation then deletion | 0.0085 | 0.0060 | 0.014 |
| Multiple imputation | 0.0026 | 0.022 | 0.024 |
| Multiple imputation then deletion | 0.0078 | 0.0059 | 0.014 |
| **MNAR - scenario 3** | | | |
| Complete case analysis | 0.0076 | 0.031 | 0.039 |
| Single imputation | 0.0084 | 0.023 | 0.031 |
| Single imputation then deletion | 0.0059 | 0.019 | 0.025 |
| Multiple imputation | 0.0017 | 0.033 | 0.034 |
| Multiple imputation then deletion | 0.0053 | 0.017 | 0.022 |

**Supplementary Table 3.** Results from the methods for handling missing data in a dichotomous outcome

MCAR: Missing completely at random

MAR: Missing at random

MNAR: Missing not at random

Scenario 1: 10% of data missing in both case-mix (age, sex, and baseline NIHSS) and outcome (mRS) in all 17 hospitals.

Scenario 2: 40% of data missing in the outcome and baseline NIHSS equally distributed over all hospitals.

Scenario 3: 40% of data missing in the outcome and baseline NIHSS with the probability of missing data varying between hospitals.

| | **Mean Variance** | **Mean Squared Bias Error** | **Mean Squared Error** |
|---|---|---|---|
| **MCAR - scenario 1** | | | |
| Complete case analysis | 0.0038 | 0.00028 | 0.0041 |
| Single imputation | 0.0032 | 0.0028 | 0.0060 |
| Single imputation then deletion | 0.0021 | 0.00040 | 0.0025 |
| Multiple imputation | 0.0015 | 0.0028 | 0.0042 |
| Multiple imputation then deletion | 0.0018 | 0.00064 | 0.0024 |
| **MAR - scenario 1** | | | |
| Complete case analysis | 0.0040 | 0.00032 | 0.0043 |
| Single imputation | 0.0029 | 0.0026 | 0.0055 |
| Single imputation then deletion | 0.0020 | 0.00032 | 0.0023 |
| Multiple imputation | 0.0014 | 0.0024 | 0.0038 |
| Multiple imputation then deletion | 0.0017 | 0.00055 | 0.0022 |
| **MNAR - scenario 1** | | | |
| Complete case analysis | 0.0039 | 0.00044 | 0.0043 |
| Single imputation | 0.0034 | 0.0026 | 0.0060 |
| Single imputation then deletion | 0.0024 | 0.00044 | 0.0029 |
| Multiple imputation | 0.0017 | 0.0029 | 0.0046 |
| Multiple imputation then deletion | 0.0021 | 0.00064 | 0.0028 |
| **MCAR - scenario 2** | | | |
| Complete case analysis | 0.014 | 0.0046 | 0.019 |
| Single imputation | 0.0045 | 0.026 | 0.030 |
| Single imputation then deletion | 0.0078 | 0.0052 | 0.013 |
| Multiple imputation | 0.0030 | 0.022 | 0.025 |
| Multiple imputation then deletion | 0.0072 | 0.0056 | 0.013 |

| | | | |
|---|---|---|---|
| **MAR - scenario 2** | | | |
| Complete case analysis | 0.014 | 0.0048 | 0.018 |
| Single imputation | 0.0038 | 0.026 | 0.030 |
| Single imputation then deletion | 0.0065 | 0.0057 | 0.012 |
| Multiple imputation | 0.0027 | 0.021 | 0.024 |
| Multiple imputation then deletion | 0.0059 | 0.0058 | 0.012 |
| **MNAR - scenario 2** | | | |
| Complete case analysis | 0.016 | 0.0059 | 0.022 |
| Single imputation | 0.0045 | 0.026 | 0.031 |
| Single imputation then deletion | 0.0098 | 0.0046 | 0.014 |
| Multiple imputation | 0.0033 | 0.022 | 0.025 |
| Multiple imputation then deletion | 0.0087 | 0.0052 | 0.014 |
| **MCAR - scenario 3** | | | |
| Complete case analysis | 0.013 | 0.0046 | 0.018 |
| Single imputation | 0.0047 | 0.025 | 0.029 |
| Single imputation then deletion | 0.0071 | 0.0050 | 0.012 |
| Multiple imputation | 0.0030 | 0.021 | 0.024 |
| Multiple imputation then deletion | 0.0066 | 0.0053 | 0.012 |
| **MAR - scenario 3** | | | |
| Complete case analysis | 0.012 | 0.0092 | 0.022 |
| Single imputation | 0.0038 | 0.027 | 0.031 |
| Single imputation then deletion | 0.0073 | 0.0077 | 0.015 |
| Multiple imputation | 0.0029 | 0.024 | 0.027 |
| Multiple imputation then deletion | 0.0065 | 0.0078 | 0.014 |
| **MNAR - scenario 3** | | | |
| Complete case analysis | 0.013 | 0.017 | 0.030 |
| Single imputation | 0.0042 | 0.028 | 0.033 |
| Single imputation then deletion | 0.0086 | 0.0076 | 0.016 |
| Multiple imputation | 0.0033 | 0.025 | 0.028 |
| Multiple imputation then deletion | 0.0077 | 0.0076 | 0.015 |

### Supplementary Figures

**Supplementary Figure 1.** Mean squared error versus reference centre effect per method for each scenario under (A) MCAR, (B) MAR, and (C) MNAR when analysing an ordinal outcome

MCAR: Missing completely at random

MAR: Missing at random

MNAR: Missing not at random

Scenario 1: 10% of data missing in both case-mix (age, sex, and baseline NIHSS) and outcome (mRS) in all 17 hospitals.

Scenario 2: 40% of data missing in the outcome and baseline NIHSS equally distributed over all hospitals.

Scenario 3: 40% of data missing in the outcome and baseline NIHSS with the probability of missing data varying between hospitals.

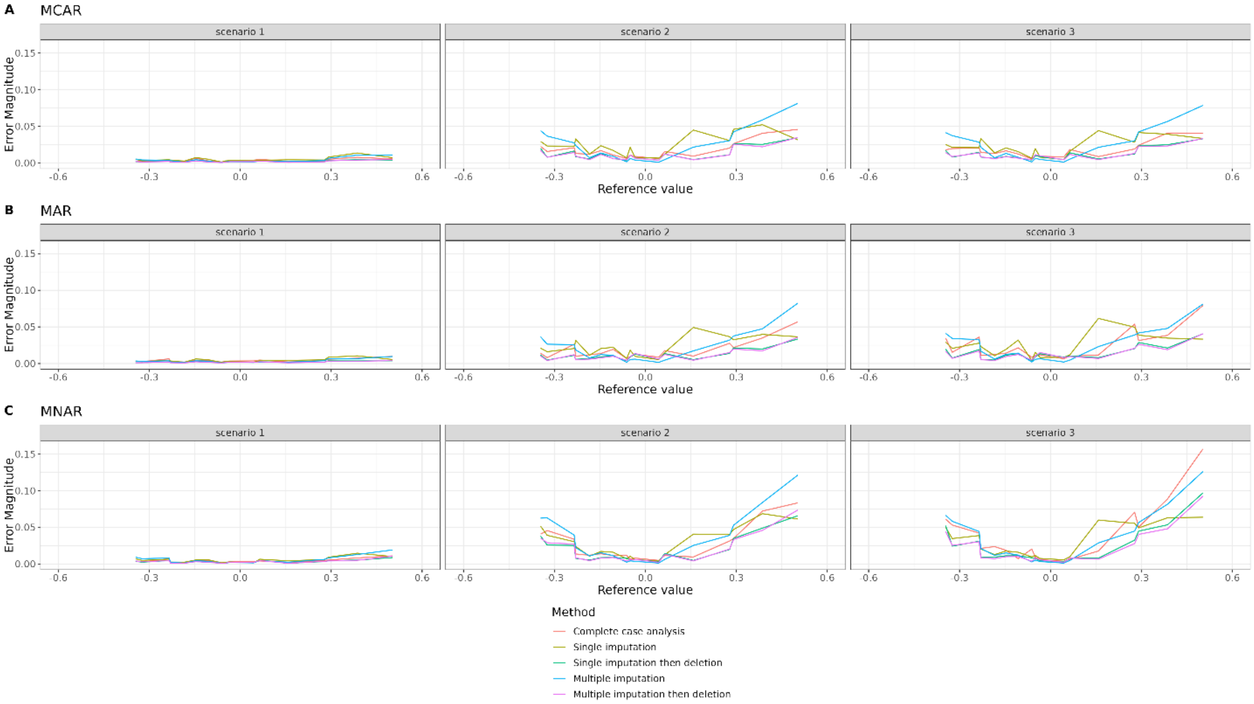

**Supplementary Figure 2.** Mean squared error versus reference centre effect per method for each scenario under (A) MCAR, (B) MAR, and (C) MNAR when analysing a dichotomous outcome

MCAR: Missing completely at random

MAR: Missing at random

MNAR: Missing not at random

Scenario 1: 10% of data missing in both case-mix (age, sex, and baseline NIHSS) and outcome (mRS) in all 17 hospitals.

Scenario 2: 40% of data missing in the outcome and baseline NIHSS equally distributed over all hospitals.

Scenario 3: 40% of data missing in the outcome and baseline NIHSS with the probability of missing data varying between hospitals.

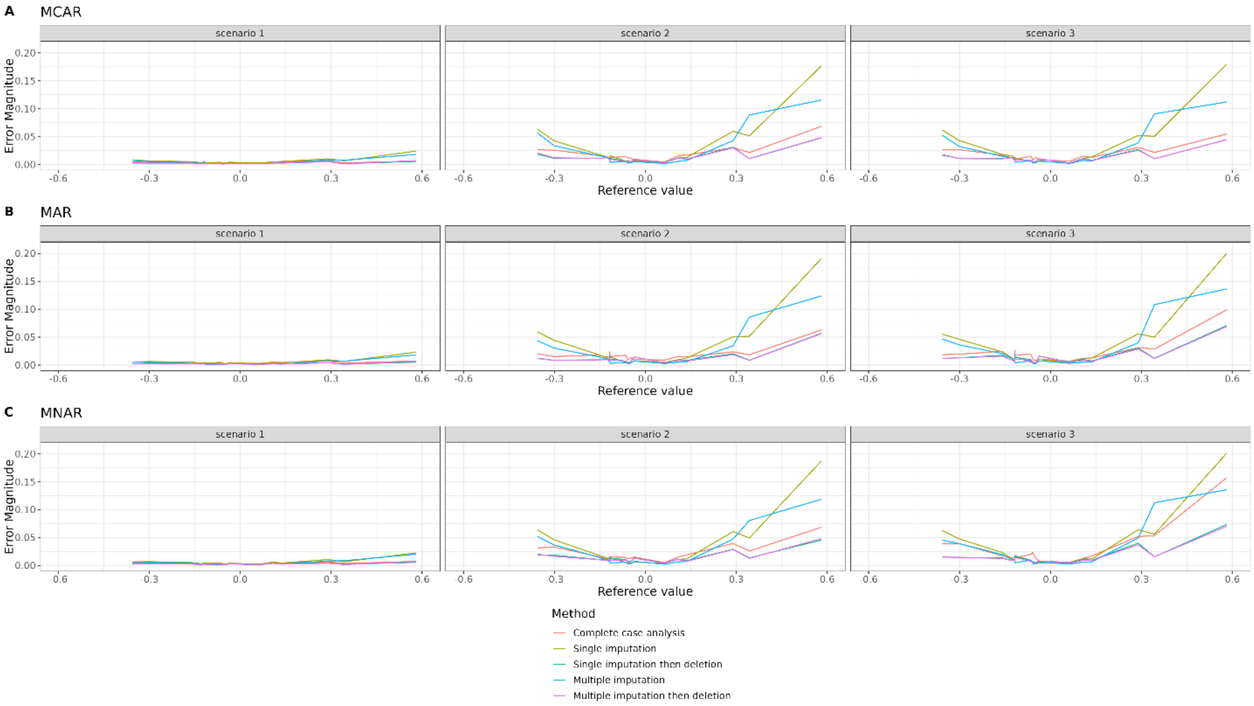
